# Diagnostic performance of the faecal immunochemical test for patients with low-risk symptoms of colorectal cancer in primary care: a service evaluation in the South West of England

**DOI:** 10.1101/2020.08.21.20173534

**Authors:** Sarah ER Bailey, Gary A Abel, Alex Atkins, Rachel Byford, Sarah-Jane Davies, Joe Mays, Timothy J McDonald, Jon Miller, Catherine Neck, John Renninson, Paul Thomas, Fiona M Walter, Sarah Warren, Willie Hamilton

## Abstract

**Objectives:** To evaluate the faecal immunochemical test (FIT) for primary care clinicians to triage patients with low-risk symptoms of possible colorectal cancer, and to estimate its diagnostic performance.

**Design:** Service delivery evaluation.

**Setting:** All primary and secondary care providers in the South West of England, approximate population 4 million.

**Participants:** 3890 patients aged ≥50 years presenting in primary care with low-risk symptoms of colorectal cancer, following NICE NG12 and DG30, with a FIT (HM-JACKarc assay) analysed from 01/06/2018 to 31/12/2018.

**Main outcome measures:** Diagnosis of colorectal cancer.

**Results:** 618 (15.9%) patients tested positive at a threshold of 10μg Hb/g faeces (median 36μg Hb/g faeces (IQR 17 to 149)); 458 (74.1%) of these had an urgent referral to specialist lower gastrointestinal (GI) services within three months. 43 were diagnosed with colorectal cancer within 12 months. 3272 patients tested negative; 324 (9.9%) were referred on an urgent lower GI pathway in secondary care within three months. 8 were diagnosed with colorectal cancer within 12 months. The positive predictive value of FIT for colorectal cancer in the low-risk symptomatic population was 7.0% (95% CI 5.1% to 9.3%) and the negative predictive value was 99.8% (CI 99.5% to 99.9%). Sensitivity was 84.3% (CI 71.4% to 93.0%),and specificity 85.0% (CI 83.8% to 86.1%). The area under the ROC curve was 0.92 (CI 0.86 to 0.96). A threshold of 37μg Hb/g faeces would identify patients with an individual 3% risk of cancer.

**Conclusions:** FIT performs exceptionally well to triage patients with low-risk symptoms of colorectal cancer in primary care. The threshold value of 10μg Hb/g faeces represents a risk of cancer below 3% used in current NICE guidance; however, this lower value may be appropriate to meet the national aspiration of improving cancer diagnostics.

## Introduction

There are around 1.8 million new colorectal cancer diagnoses worldwide each year, and almost 900,000 deaths.^1^ Population screening is effective in reducing mortality, with a relative risk of colorectal cancer mortality varying between 0.67 and 0.88 depending upon the screening modality, frequency of screening, and sex.^2^ However, even when screening is available, most colorectal cancers present with symptoms. In the UK, less than 10% of colorectal cancers are identified by screening, with the remainder identified after symptoms have developed.^3^ In many countries, symptomatic patients present first to primary care, where the general practitioner (GP) assesses the possibility of cancer, and investigates or refers for specialist tests if appropriate.^4^ The usual diagnostic test in secondary care is colonoscopy, with CT imaging or capsule endoscopy occasionally used.

Requests for urgent colorectal cancer investigation have relentlessly increased over the last decade, with a parallel increase in colonoscopies. These doubled in the UK between 2012 and 2017.^5^ This rise was driven in part by referral of patients whose symptom profile, whilst still representing possible cancer, was relatively low-risk.^6^ These patients, often with abdominal pain or mild anaemia, had been excluded from UK national guidance in 2005,^7^ but transpired to have the worst survival across the different symptoms, often presenting as an emergency.^8 9^ In 2015, the National Institute for Health and Care Excellence (NICE) published revised guidance, NG12.^10^ The revised NICE recommendations were explicitly based on the risk of cancer posed by the patient’s symptoms, and used only primary care evidence to estimate this risk. Patients having a risk of colorectal cancer of 3% or more are recommended for an urgent suspected cancer referral, and are usually offered colonoscopy. For risks below 3%, patients were to be offered testing for occult blood in their faeces, with those testing positive to be referred urgently. This recommendation was based on a systematic review performed by NICE, finding six studies of faecal occult blood testing mostly in secondary care, totalling 9871 patients.^10^ The sensitivity and specificity for colorectal cancer varied considerably across these studies, though the diagnostic performance was considered sufficient in the absence of other tests available in primary care. An economic evaluation supported this recommendation.^10^

Faecal immunochemical test (FIT) for haemoglobin measure the amount of haemoglobin in a stool sample and have largely replaced faecal occult blood testing in screening programmes. FITs are considered to be superior to faecal occult blood tests; they require only one stool sample, are not sensitive to dietary factors or medications, provide a fully quantitative result, and have higher uptake rates, so appear more acceptable to patients. Further NICE guidance issued in 2017 (DG30) recommended FIT should replace faecal occult blood testing in primary care patients with low-risk symptoms of colorectal cancer.^11^ The systematic review underpinning that recommendation found nine studies:^12^ in only one was the FIT performed in primary care, though even in that study all patients had already been selected for urgent referral for possible colorectal cancer.^13^ Thus all the evidence underpinning the use of FITs in primary care in DG30 was from the high-risk referred population; this brings a substantial risk of spectrum bias.^14^

This study evaluated the FIT service provided to GPs for patients with low-risk symptoms of possible colorectal cancer in the South West of England, and estimated the diagnostic performance of FITs in this community.

## Methods

This joint South West Cancer Alliances transformation project provided a quantitative faecal immunochemical test (FIT) service to primary care practices across the South West of England (population approx. 4 million) from June 2018. This area includes 14 secondary care providers and 10 clinical commissioning groups (CCGs), listed in Appendix 1.

The FIT diagnostic service (comprising of FIT test kits for patients, patient instructions, lab processing of FIT and timely associated reporting of results) was available to GPs to triage patients with low-risk symptoms of colorectal cancer, as defined by NG12 and DG30.^10 11^ Patients meeting the following criteria were eligible for a FIT test (these criteria were derived from the original NG12 for faecal occult blood testing, since removed from the guidance but current at the time of project design):

- Aged 50 years and over with unexplained abdominal pain or weight loss
- Aged 50 to 60 years with change in bowel habit or iron deficiency anaemia
- Aged 60 years and over with anaemia, even in the absence of iron deficiency

The laboratory service was provided by Severn Pathology in Bristol and the Exeter Clinical Laboratory in Exeter using the HM-JACKarc analyser. This assay has a recommended analytical range of 7 to 400μg Hb/g faeces (though some values below 7μg Hb/g faeces were reported); results over 400μg Hb/g faeces were recorded as >400μg Hb/g faeces. A threshold value of ≥10μg Hb/g faeces defined a ‘positive’ test, as per DG30.^11^ Test kit packs were delivered to primary care practices including: the test unit, instructions for use, a form to select the indication for the test, and a pre-paid envelope for the patient to return the completed test to the laboratory. Test results were returned to practices electronically in Devon, Cornwall, and Avon. In Somerset, Wiltshire, and Gloucestershire, reports were initially sent by post, and later electronically.

Information about the service was publicised through local CCG newsletters and through the local Cancer Research UK Facilitator Team, who provided practice-level training and support. GPs were provided with written, online, and video support for using the FIT service, indications for the test and how to use it, and advice on how to deal with a positive test. GPs were advised in the guidance that if FIT was positive they should consider using an urgent referral for suspected cancer under the local secondary care provider’s arrangements. They were also advised that occult blood in the stool can be caused by a wide variety of benign conditions as well as colorectal cancer, and further assessment may be appropriate to rule these out before referring.

### Data collection

All patients with a FIT analysed from 1^st^ June 2018 to 31^st^ December 2018 were included in this study. Data extracted from the two laboratories included the test date, result, indication, patient year of birth, and gender. Separately, each of the 14 secondary care providers in the region extracted data, including stage at diagnosis, on any cancer identified from 1^st^ June 2018 to 31^st^ December 2019 after entry into upper or lower gastro-intestinal services. This captured cancers diagnosed by all routes, including screening and incidental findings such as routine referral or emergency admission. This allowed for 12 months of follow-up time for all patients, during which any missed colorectal cancer diagnoses in FIT negative patients were very likely to be diagnosed through other routes. Cancers diagnosed in FIT negative patients after 12 months may not be deemed to have been “missed” as it is improbable they would have been present and causing symptoms at the time of testing. Only cancers identifiable on a gastro-intestinal pathway were identified: non-GI cancers, referred to other cancer diagnostic pathways, were not identified.

FIT results were matched against referral and diagnosis data by each of the secondary care providers using the NHS number, with this NHS number removed and replaced with a randomly allocated study number. Year of birth was used as a secondary confirmation of correct matching of patient records. This was done to adhere with information governance requirements and to ensure completeness of the full patient pathway. GPs were advised not to offer multiple FITs to individual patients; where more than one test was recorded for one patient, the earliest test result was used.

### Statistical analysis and power calculation

Summary statistics were used to describe the cohort, and to estimate the performance of FIT in this population, including sensitivity, specificity, positive predictive value (PPV), and negative predictive value. A chi-squared test was used to compare the proportion of male participants, and a Mann-Whitney test to compare the median age, between those with a positive and those with a negative test result. A receiver-operating characteristic curve was produced for the quantitative FIT result against colorectal cancer diagnosis. Logistic regression was used to model the relationship between cancer and FIT, with FIT results treated as a continuous variable, after log-transformation to improve the final model fit. Non-linearity in the relationship between FIT result and cancer was explored using fractional polynomials, though goodness of fit was not improved by doing so. Consequently, a linear term was retained. The probability of being diagnosed with cancer in the next year for a given FIT value was estimated from the final model, in particular identifying the value equating to an individual cancer risk of 3%, to mirror NICE recommendations for urgent investigation.^10^ Stata version 16 was used for all analyses.^15^ Diagnostic test summary statistics were estimated with the DIAGT module.^16^

A simulation approach was used to estimate the sample size required to achieve 95% confidence intervals of 2.2% to 4% around a cancer risk of 3% from the logistic regression. Assuming a linear relationship between the FIT result and cancer risk suggested a sample size of 2250 would be sufficient so long as the FIT threshold was within the central core of the distribution of FIT levels. It was estimated that 10,000 tests would be used in a year; data were collected over seven months to meet the sample size requirement. In practice it was comfortably exceeded, increasing precision.

### Data governance

As this project was evaluating service delivery, and not changing routine clinical practice, ethical approval was not required. Data sharing agreements were drawn up between all parties, and Caldicott guardian approvals were in place to allow data sharing. The requirement for individual NHS numbers for use within this evaluation meets the criteria set out in section 6 of the General Data Protection Regulation: Guidance on Lawful Processing. The processing of data is based upon GDPR Article 6(1)(e) – ‘exercise of official authority’ and article 9(2)(h) ‘management of health and care services’. The enabling legislation is the NHS Act 2006 section 13E, including the duty on NHS England to ‘secure continuous improvement in the quality of services’. The same basis supported the secondary care providers supplying data. The study protocol is available on the University of Exeter website at http://hdl.handle.net/10871/122303.

## Results

From 1^st^ June 2018 to 31^st^ December 2018, 3,890 FIT samples were submitted to and analysed by the two laboratories. The median age of tested patients was 65 years (interquartile range (IQR): 56 to 75) and 1644 (42.6%) were male. Criteria for investigation were: 1617 (41.6%) aged ≥50 years with abdominal pain or weight loss; 1194 (30.7%) aged <60 years with changes in bowel habit or iron deficiency anaemia; 930 (23.9%) aged >60 years and with anaemia (in absence of iron deficiency). No criteria for investigation were recorded in 149 (3.8%).

### FIT results

A faecal haemoglobin result of ≥10μg Hb/g faeces (test positive) was recorded for 618 patients (15.9%). Patients testing positive were more often male (46.1% vs 42.0%, p=0.017) and older (median age 71 vs 63, IQR 60 to 79, p<0.001). Of patients testing positive, the median age was 71.7 years (IQR 60.1-79.7); 288 (46.3%) were male. The median result in FIT positive patients was 36μg Hb/g faeces (IQR 17 to 149). Figure 1 shows the distribution of FIT results in patients with a positive result.

**Figure 1:**
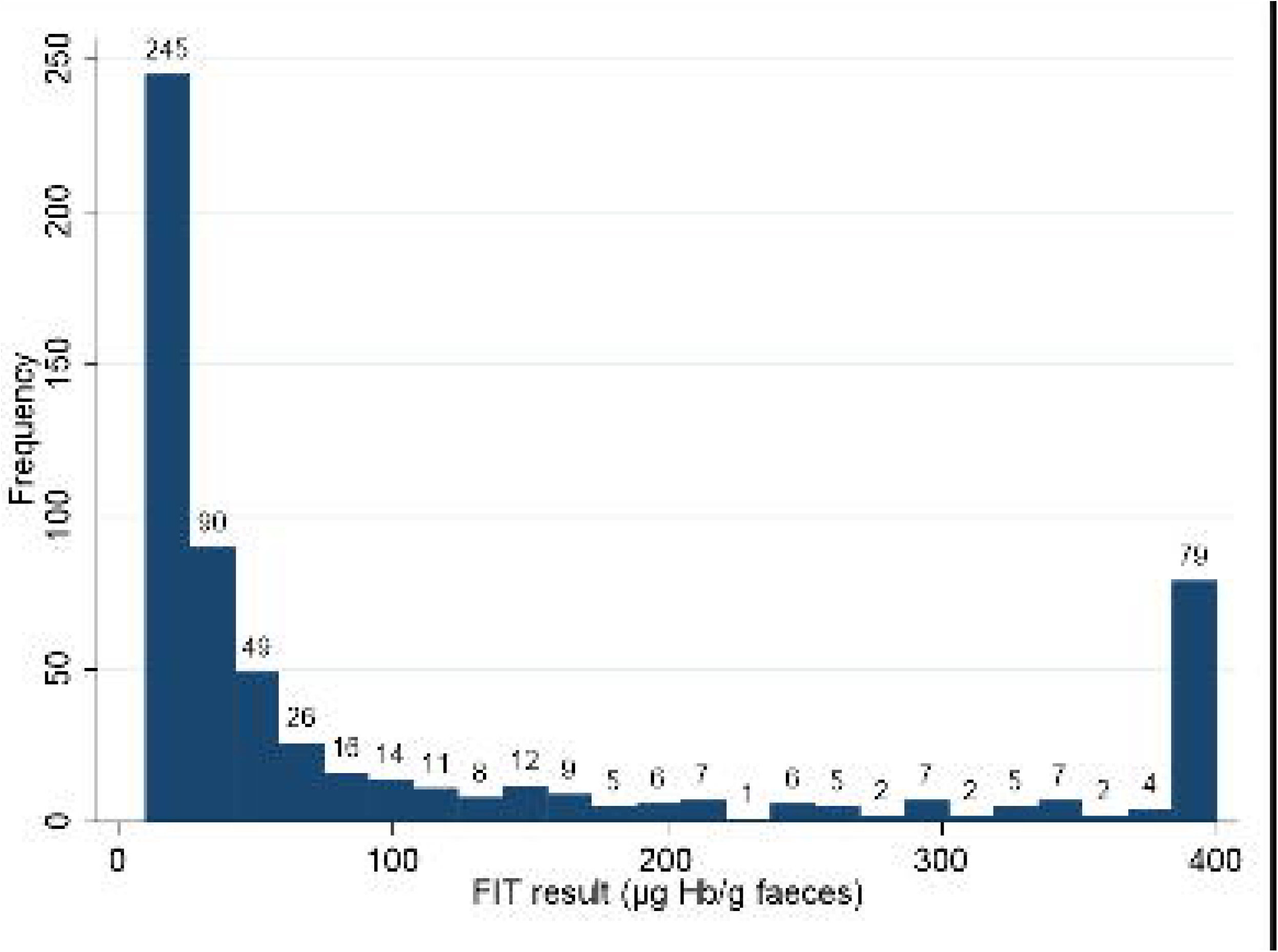
Histogram of positive FIT results in 618 primary care patients, with low-risk symptoms of possible colorectal cancer.

### Referrals in patients with a FIT

Gastrointestinal (GI) referrals and cancer outcomes are summarised in Figure 2. This shows lower GI referrals within 93 days of the positive FIT test, and cancer outcomes within 12 months.

**Figure 2:**
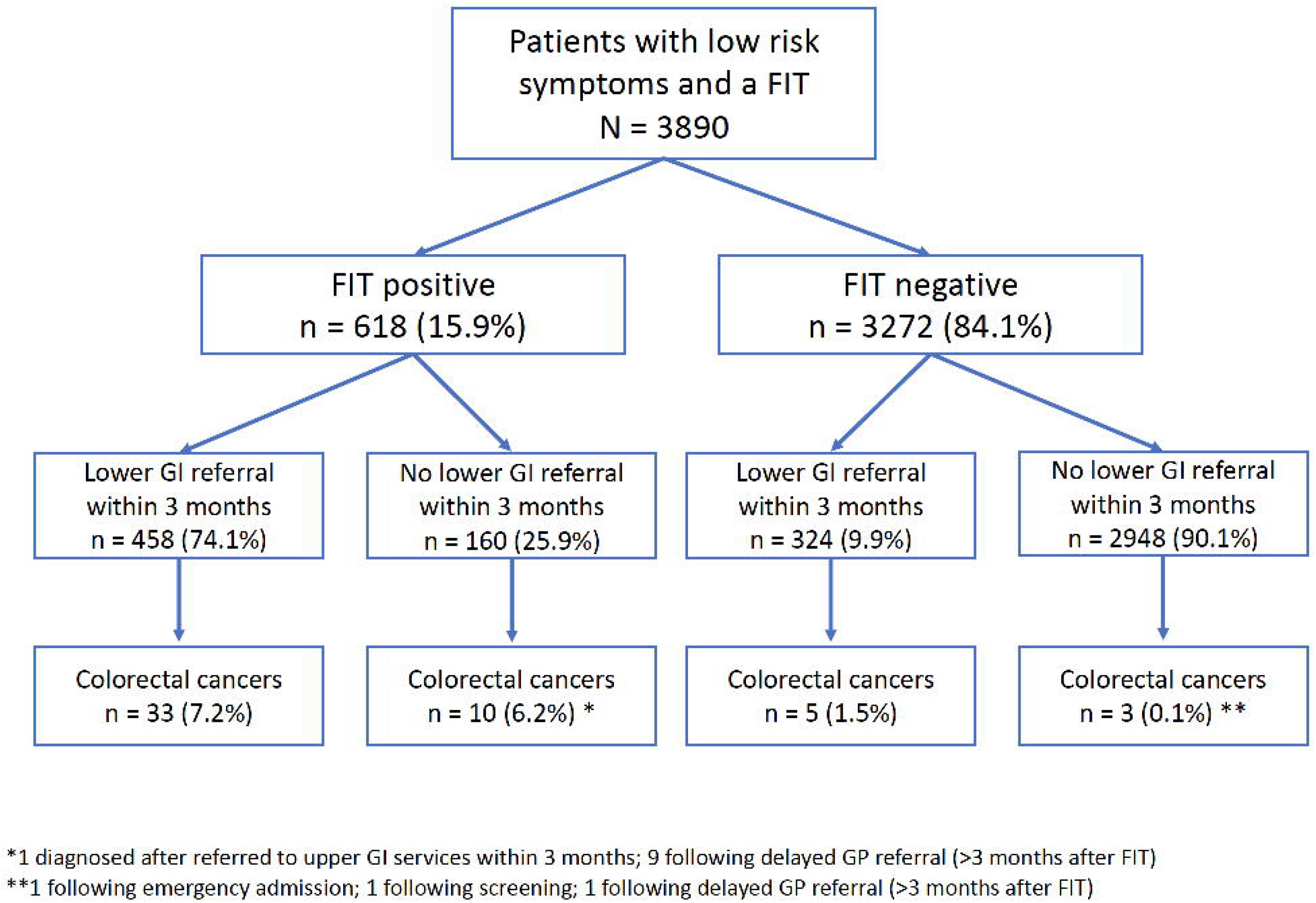
flow diagram to show number of patients tested, test results, GI referrals and colorectal cancers.

### Cancer outcomes

Table 1 shows the cancers identified during the year after FIT. The positive predictive value of FIT in this low-risk symptomatic population is 7.0% (95% CI 5.1% to 9.3%), and the negative predictive value is 99.8% (CI 99.5% to 99.9%). The sensitivity in this population is 84.3% (CI 71.4% to 93.0%), and the specificity 85.0% (83.8% to 86.1%). The area under the ROC curve is 0.92 (CI 0.86 to 0.96) (Figure 3).

**Table 1:** the number of colorectal, oesophago-gastric, and pancreatic cancers diagnosed in total, and by FIT result, within 12 months of FIT date.

| Cancer | Total | ≥10µg Hb/g faeces<br>N=618 | <10µg Hb/g faeces<br>N=3264 |
| --- | --- | --- | --- |
| Colorectal | 51 | 43 (7.0%) | 8 (0.2%) |
| Oesophago-gastric | 9 | 3 (0.5%) | 6 (0.2%) |
| Pancreatic | 7 | 1 (0.2%) | 6 (.02%) |
| Total | 67 | 47 (7.6%) | 20 (0.6%) |

**Figure 3:**
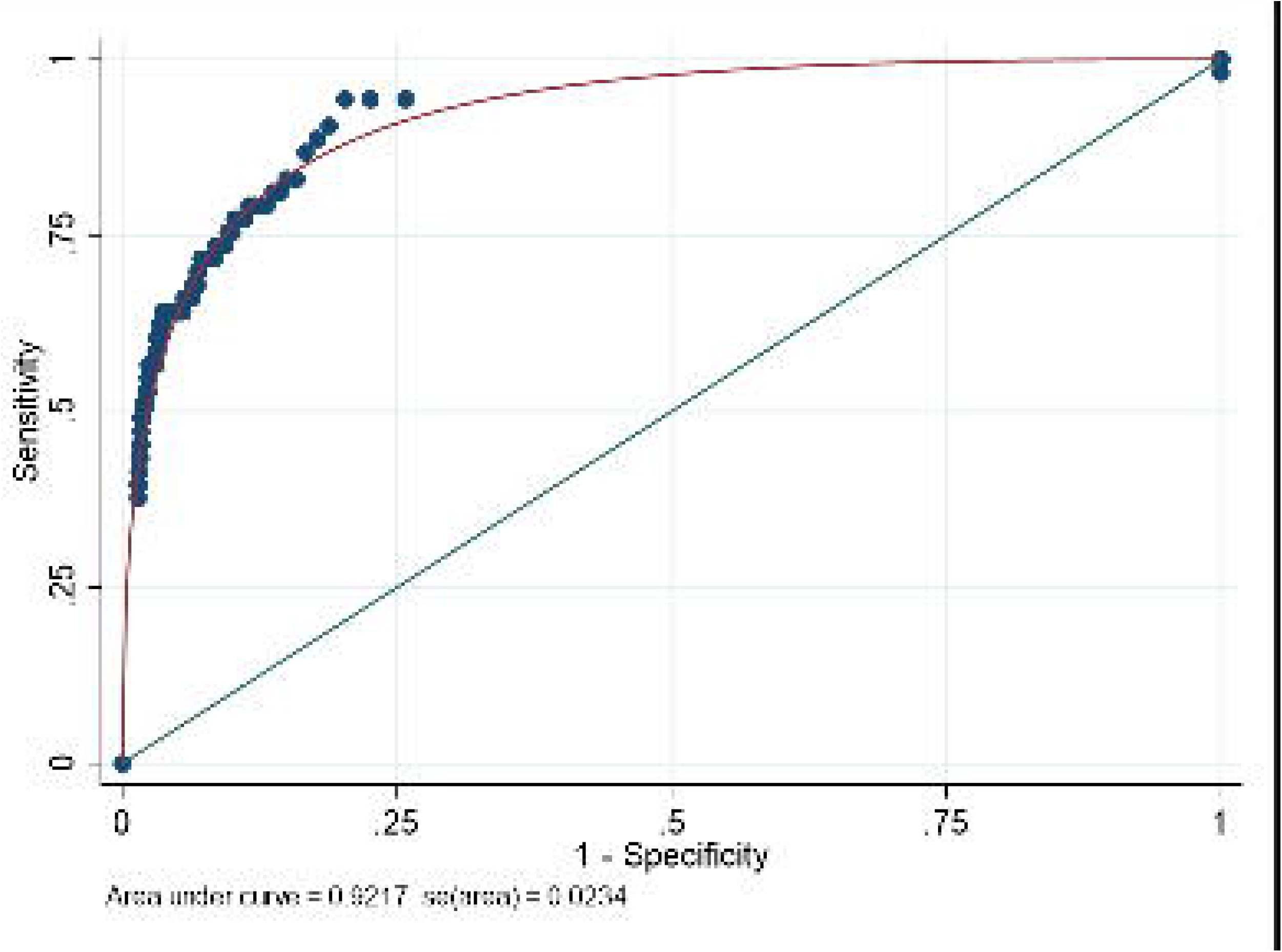
receiver operator characteristic (ROC) curve for the faecal immunochemical test (FIT) in the low-risk symptomatic primary care population.

The median number of days from FIT to diagnosis of colorectal cancer in patients with a positive FIT was 34 (IQR 23 to 56). Staging data were available for 31 of 43 patients: 6 Dukes’ A; 5 Duke’s B; 12 Dukes’ C; 8 Dukes’ D. The median number of days to diagnosis in patients with a negative FIT was 57 days (IQR 37 to 197). Staging data were available for six of eight patients: 1 Duke’s B; 2 Dukes’ C; 3 Dukes’ D.

### Cancer risk by FIT result

Figure 4 shows the estimated probability that an individual will be diagnosed with colorectal cancer for a given FIT result, estimated from the logistic regression model. Using this model, a FIT result of 37μg Hb/g faeces (CI 26 to 50) in an individual with that result corresponds to a colorectal cancer risk of 3%.

**Figure 4:**
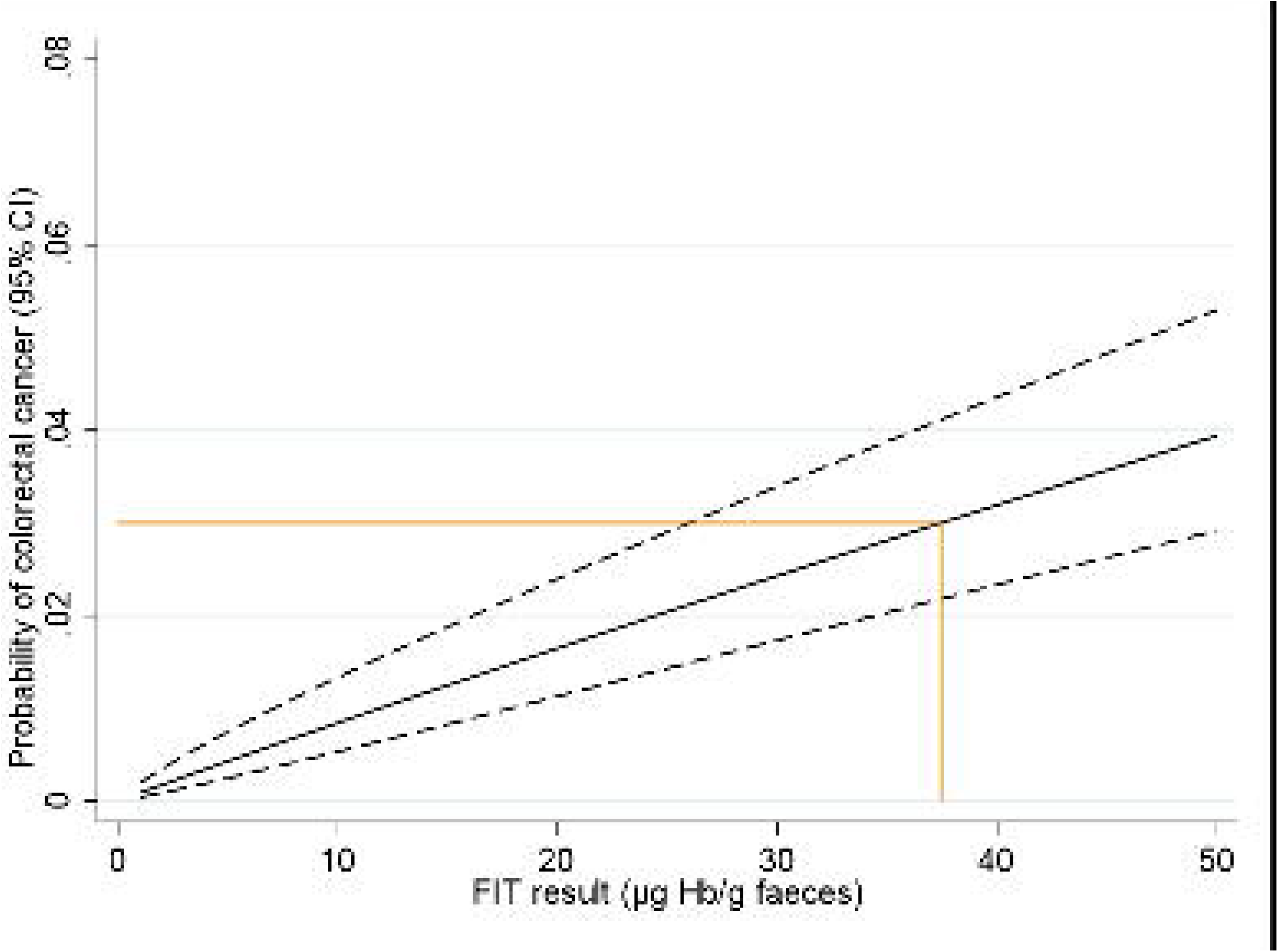
predicted probability of colorectal cancer against FIT result, with 95% confidence limits. The orange line shows the FIT result that corresponds to a colorectal cancer risk of 3% in an individual with that result.

## Discussion

This study reports the use of FIT for detection of colorectal cancer in a primary care symptomatic population. The test performed very well using the threshold value of ≥10μg Hb/g faeces. Test sensitivity and specificity were 84.3% and 85.0% respectively, both notably high figures for a primary care cancer test. Using this threshold, the positive predictive value of a positive FIT result was 7.0%, and the negative predictive value 99.8%, in a population with an overall prevalence of colorectal cancer of 1.3%. The FIT test also performed well irrespective of gender or age. A FIT result of 37μg Hb/g faeces corresponded to an individual’s colorectal cancer risk of 3%.

### Strengths and limitations

The strengths of this study are its size, and its setting being where the test will be used, eliminating spectrum bias. The three symptom groupings used by GPs to prompt FIT testing were estimated to have PPVs in primary care in the range 1-3%, and the overall prevalence of 1.3% fell within that range. Every one of the 14 secondary care providers in the region were recruited, increasing reliability and generalisability. Cancer metrics in the NHS are very accurately maintained; dedicated cancer managers ensure accurate data recording, and secondary care provider performance on cancer metrics is regularly published in the public domain. Furthermore, all secondary care providers of cancer services within England are required to use nationally defined datasets eliminating disparity in data definitions. Despite the thorough methods, it is possible that a small number of cancers were missed, though this is unlikely to affect the overall interpretation of the results. Crucially, the methods allowed the identification of colorectal cancers in those not offered further investigation after the FIT result was received, and a long follow-up period of one year was achieved.

The age group studied, with a median age of 65 years for those tested, is close to the median age for colorectal cancer diagnosis of 72 years, suggesting the GPs were using the test in those genuinely considered to have a real - but small - risk of cancer. More women were tested, whereas colorectal cancer is slightly more common in men. This may reflect the entry criteria, particularly with two of the three criteria incorporating iron-deficiency anaemia, a condition more common in women,^17^ or the fact that women are more likely to seek medical intervention.^18^ Symptom data could not be verified, but the overall prevalence figure suggests testing was rarely extended into higher-risk groups. Completing the test was patient driven; only tests which were completed and returned to the lab were reported; it is not known how many tests were handed out by GPs and not returned to the labs.

### Comparison with previous literature

Two recent studies can be compared, as both examined FIT in the symptomatic primary care population, rather than the screening or referred populations: both also used 10[ig Hb/g faeces to define a positive test. Juul *et al* studied 3462 Danish primary care patients aged ≥30 years, with symptoms not meriting urgent colonoscopy, but not defined further.^19^ In that study, FIT was also recommended in patients diagnosed with irritable bowel syndrome, lest this were a misdiagnosis. FITs were positive in 15.6%, and 9.4% of these (CI 7.0 to 11.9%) had a colorectal cancer diagnosed in the next three months. There were fewer than three cancers identified in FIT negative patients (the inexact number reflecting Danish data protection rules). Nicholson *et al* followed up 9896 primary care patients in Oxfordshire, England, for six months after FITs were performed in primary care. The entry criteria did not match NICE guidance DG30 or NG12, and included rectal bleeding. The sensitivity for colorectal cancer was 90.5% (CI 84.9-96.1%), and the positive predictive value of a positive test 10.1% (CI 8.2-12.0%).^20^ The PPV of a positive result in the present study of 7% is the lowest of the three, which may reflect the stricter criteria for use, in particular excluding patients with rectal bleeding, but matching current national guidance.^10^ As a comparison, the sensitivity of 84.3% reported here is higher than in the screening population of 67.0% (CI 59.0% to 74.0%, with thresholds of >50 ng/g^21^), and lower than that in referred populations of 93.3% (CI 80.7% to 98.3%, thresholds of >10 ng/g).^22^

### Clinical and research use of the results

The values reported in this study are excellent for a cancer triage test in primary care. The performance of diagnostic tests is generally worse as the prevalence of the target condition falls.^14^ In primary care, gastrointestinal complaints are common, and the symptoms of colorectal cancer overlap both with less common cancers, such as pancreatic or oesophago-gastric, and with common benign conditions. With most symptoms (apart from rectal bleeding) the likelihood of colorectal cancer is low, often in the range 1-3%.^23^ A large UK primary care study showed that patients would opt for cancer investigation even for risks as low as 1%.^24^ FIT testing has been introduced to allow primary care investigation of such patients. It works in classical Bayesian fashion: from a prior risk of colorectal cancer of 1.3% in the symptomatic population, a positive test increased the risk to 7.0%, and a negative test reduced it to 0.2%, which is approximately the whole population background risk, including those without symptoms.^23^ Furthermore, in five of the eight cancers with FIT negative results, the patient’s GP still requested urgent investigation for possible colorectal cancer, probably because continuing symptoms allowed the GP to ‘overrule’ the negative test. Conversely, nearly a quarter of patients with a positive FIT test were not offered investigation within three months, though ultimately all who had colorectal cancer were investigated within a year.

Other cancers were diagnosed in participants in this study. Sixteen oesophago-gastric or pancreatic cancers were found, only four having a positive FIT test. This suggests that GPs should consider other intra-abdominal cancers in patients with a negative FIT and continuing symptoms.

Whilst the PPV of a positive test at or above the current threshold of 10μg Hb/g faeces is 7%, the risk of having colorectal cancer for an individual with a FIT result of exactly this value is 1% or lower (Figure 4). Given this risk is lower than the 3% chosen to underpin the NICE NG12 recommendations for urgent cancer investigation,^10^ there may be scope to raise the threshold at which urgent definitive investigations are undertaken. A FIT result of 37μg Hb/g faeces (CI 26 to 50) would identify those with a personal 3% risk of cancer, though the large uncertainty on this estimate may warrant the use of a lower value until more data are available to reduce this uncertainty. Such a change may be appropriate while endoscopy resources are severely curtailed by COVID-19 precautions, with ‘safety netting’ by GP review for those with results between 10 and 36μg Hb/g.faeces.^25^ In the long term however, the UK’s aspiration is for improvements in cancer diagnosis to increase the proportion of cancer patients diagnosed at stage I or II to 75% by 2028 (from a pre-COVID approximate 53%).^26^ If colorectal cancer improvements are to contribute to this target, it may be that the threshold should be retained at 10μg Hb/g faeces, or even lowered further, though not below the level where the test is considered reliable, currently 7μg Hb/g faeces.

Several research needs arise from this study. The first is a health-economic analysis, examining the choice of FIT threshold from that perspective. Second, it may be possible to combine data on symptoms and demographics with the FIT result to increase the predictive power of FIT. A third strand of research – not directly related to this study, but overlapping – considers whether FIT can be used to triage the high-risk population. Such studies are underway, with particular urgency in the post-COVID recovery phase. They will complement the study reported here, and establish the final place for FIT in colorectal cancer triage.

## Conclusion

FIT testing in the low-risk primary care population performs well. False-negatives are few in number, and many of those with a false-negative test appear to receive timely investigation despite the negative test. The false-positive rate is 93%, meaning 13 patients with a positive FIT have to undergo colonoscopy to identify one cancer. This is a major diagnostic advance; low-risk patients were previously either not investigated, and had more emergency admissions and worst survival,^8, 9^ or were referred for colonoscopy. The background rate of cancer of 1.3% in this population meant 77 patients had to be offered colonoscopy to identify one cancer, potentially swamping endoscopy services, and putting patients at a small risk of complications. Clinically, therefore, FIT works, though health-economic aspects are as yet uncertain.

## Data Availability

As the raw data analysed in this study include patient identifiable information, we cannot share them currently. We are happy to discuss requests for data with the Caldicott guardian (JR).

## Copyright statement

The Corresponding Author has the right to grant on behalf of all authors and does grant on behalf of all authors, a worldwide licence to the Publishers and its licensees in perpetuity, in all forms, formats and media (whether known now or created in the future), to i) publish, reproduce, distribute, display and store the Contribution, ii) translate the Contribution into other languages, create adaptations, reprints, include within collections and create summaries, extracts and/or, abstracts of the Contribution, iii) create any other derivative work(s) based on the Contribution, iv) to exploit all subsidiary rights in the Contribution, v) the inclusion of electronic links from the Contribution to third party material where-ever it may be located; and, vi) licence any third party to do any or all of the above.

## Competing interests

All authors have completed the ICMJE uniform disclosure form at www.icmje.org/coi_disclosure.pdf and declare: This research arises from the CanTest Collaborative, which is funded by Cancer Research UK [C8640/A23385], of which WH and FMW are Directors, GAA is Co-investigator and SB is Senior Postdoctoral Researcher. This funding source had no role in writing the manuscript or deciding to submit it for publication. It was also supported by the Peninsula Cancer Alliance, the Somerset, Wiltshire, Avon, and Gloucestershire (SWAG) Cancer Alliance, and NHS England. All authors declare: no financial relationships with any organisations that might have an interest in the submitted work in the previous three years; no other relationships or activities that could appear to have influenced the submitted work.

## Author contributions

All authors were involved in the design and inception of the study. SB, PT, TMcD, AA, SJD, CN, SW, RB, JM, and JR contributed to data collection and data governance. SB, GA, and WH carried out data analysis and created the first draft of the manuscript along with FW. All authors reviewed and contributed to the final draft of the manuscript. SB is responsible for the overall content as guarantor. The corresponding author attests that all listed authors meet authorship criteria and that no others meeting the criteria have been omitted.

## Transparency declaration

The lead author affirms that the manuscript is an honest, accurate, and transparent account of the study being reported; that no important aspects of the study have been omitted; and that any discrepancies from the study as originally planned have been explained.

## Ethical approval

As this project was evaluating service delivery, and not changing routine clinical practice, ethical approval was not required. Data sharing agreements were drawn up between all parties, and Caldicott guardian approvals were in place to allow data sharing. The requirement for individual NHS numbers for use within this evaluation meets the criteria set out in section 6 of the General Data Protection Regulation: Guidance on Lawful Processing. The processing of data is based upon GDPR Article 6(1)(e) - ‘exercise of official authority’ and article 9(2)(h) ‘management of health and care services’. The enabling legislation is the NHS Act 2006 section 13E, including the duty on NHS England to ‘secure continuous improvement in the quality of services’. The same basis supported the secondary care providers supplying data.

## Details of funding

This research arises from the CanTest Collaborative, which is funded by Cancer Research UK [C8640/A23385], of which WH and FMW are Directors, GAA is Co-investigator and SB is Senior Postdoctoral Researcher. It was also supported by the Peninsula Cancer Alliance, the Somerset, Wiltshire, Avon, and Gloucestershire (SWAG) Cancer Alliance, and NHS England.

## Role of funding sources

JR, JM, SJD, and JM are part of the Peninsula Cancer Alliance; their roles are described in the author contributions section. Otherwise, the funding sources had no role in the study design; in the collection, analysis, and interpretation of data; in the writing of the report; and in the decision to submit the article for publication.

## Statement of independence

The researchers were independent from the funding sources, and all authors had full access to all data in the study and can take responsibility for the integrity of the data and the accuracy of the data analysis is also required.

## Patient and public involvement statement

The study has been discussed widely within two of our research fora with public and patient representatives on several occasions. Other than that, there was no direct patient involvement.

## Acknowledgements

With thanks to Allen Barker, the cancer managers at the contributing secondary care providers, and the patients whose data contributed to this evaluation.

## Notes

### Competing Interest Statement

The authors have declared no competing interest.

### Author Declarations

The Royal Devon and Exeter NHS Foundation Trust's Health Research Authority Confidentiality Advice Team were consulted on the requirement for ethical approval for this study. They advised that we should contact individual Trusts participating in the evaluation and seek approval and confirmation from the Trust(s) Caldicott Guardian(s) that the flows of confidential patient information are legitimate and in accordance with national laws and guidelines. Each Trust involved in the evaluation was satisfied that this was the case, and that as this project was evaluating service delivery, and not changing routine clinical practice, ethical approval was not required. Data sharing agreements were drawn up between all parties, and Caldicott guardian approvals were in place to allow data sharing. The requirement for individual NHS numbers for use within this evaluation meets the criteria set out in section 6 of the General Data Protection Regulation: Guidance on Lawful Processing. The processing of data is based upon GDPR Article 6(1)(e) exercise of official authority and article 9(2)(h) management of health and care services. The enabling legislation is the NHS Act 2006 section 13E, including the duty on NHS England to secure continuous improvement in the quality of services. The same basis supported the secondary care providers supplying data.

